# COVID-19 Flow-Maps: An open geographic information system on COVID-19 and human mobility for Spain

**DOI:** 10.1101/2021.06.23.21259395

**Authors:** Miguel Ponce-de-Leon, Javier del Valle, José María Fernandez, Marc Bernardo, Davide Cirillo, Jon Sanchez-Valle, Matthew Smith, Salvador Capella-Gutierrez, Tania Gullón, Alfonso Valencia

**Author notes:** corresponding author(s): Miguel Ponce-de-Leon, Alfonso Valencia (. these authors contributed equally to this work (Miguel Ponce-de-Leon, Javier del Valle).

## Abstract

COVID-19 is an infectious disease caused by the SARS-CoV-2 virus, which has spread all over the world leading to a global pandemic. The fast progression of COVID-19 has been mainly related to the high contagion rate of the virus and the worldwide mobility of humans. In the absence of pharmacological therapies, governments from different countries have introduced several non-pharmaceutical interventions to reduce human mobility and social contact. Several studies based on Anonymised Mobile Phone Data have been published analysing the relationship between human mobility and the spread of coronavirus. However, to our knowledge, none of these data-sets integrates cross-referenced geo-localised data on human mobility and COVID-19 cases into one all-inclusive open resource. Herein we present COVID-19 Flow-Maps, a cross-referenced Geographic Information System that integrates regularly updated time-series accounting for population mobility and daily reports of COVID-19 cases in Spain at different scales of time spatial resolution. This integrated and up-to-date data-set can be used to analyse the human dynamics to guide and support the design of more effective non-pharmaceutical interventions.

## Background & Summary

COVID-19 is an infectious disease caused by SARS-CoV-2 virus, which has spread all over the world, leading to a global pandemic state. The fast progression of COVID-19 has been mainly related to two crucial factors, the high contagion rate of the virus, as well as, the worldwide mobility of humans. The high contagious rate has represented one of the leading causes of healthcare systems collapse in several countries, such as Italy and Spain, contributing to a large number of deaths. As a result, and in the absence of vaccines or other pharmacological therapies, governments from different countries have introduced several non-pharmaceutical interventions (NPI) to reduce human mobility and social contact^1^. Different NPIs include, but are not limited to, the closing of national borders, the temporal partial or total lock-downs, and social distancing measures with the aim of attenuating the epidemic^2^.

Several studies based on Anonymised Mobile (cell) Phone Data (AMPD) have been published analysing the relationship between human mobility and the spread of coronavirus^3^. For instance, in a study conducted in the USA, the authors reported a high correlation between human mobility and COVID-19 transmission rate^4^. A similar study conducted in Italy reports that mobility restrictions and social distancing policies could reduce contagions up to 45%^5^. AMPD has also been used to study how the different NPIs can change the underlying structure of the human mobility networks and how those changes affect the spread of the disease^6,7^.

AMPD has also been integrated into epidemiological models to simulate the spread of SARS-CoV-2 in time and space. For instance, a model integrating human mobility was used to predict the first wave of COVID-19 cases in Spain and to call the government to enforce a full lockdown^8,9^. Similarly, models have also been used to study the impact of mobility restrictions, physical distancing, as well as the impact of lockdown on social inequalities^10–12^. Models are critical in the design of NPIs to effectively mitigate the pandemic while reducing the negative impact on the economy, education, and other social activities^13,14^. However, developing realistic models to guide policy-makers decisions requires high quality and up-to-date data^15^. Ideally, data records should be open access, de-identified, periodically updated, and should include data on human mobility, COVID-19 daily cases, and contact tracing, among other sources of information. Furthermore, data records should be properly geo-localized with the highest possible degree of spatio-temporal resolution^16^.

Several data-sets on human mobility have been made publicly available to help analyse and model the epidemic dynamics. Pepe et al, have released a data-set based on 170,000 de-identified and aggregated smartphone users that account for movements between Italian provinces^17^. Another notable data-set includes a regularly updated multi-scale human mobility dynamic network across the United States^18^. Furthermore, data-sets on COVID-19 cases across the globe have been also released^19,20^. However, to our knowledge, none of the cited data-sets integrates cross-referenced geo-localized data on human mobility and COVID-19 cases into one all-inclusive open resource.

Herein we present a cross-referenced geographic information system (GIS) named COVID-19 Flow-Maps to manage, retrieve, visualise and analyse regularly updated time-series data on population mobility networks and daily reports of COVID-19 cases in Spain (Figure 1). Human mobility data is provided as Origin-Destination (OD) matrices at different levels of spatial and temporal resolution (district, province, and autonomous community on a daily and hourly basis). Every mobility data record reports the number of people performing zero, one, two, or more trips, on a daily basis. Health data consisting of daily confirmed COVID-19 cases is also provided on a daily basis and at different levels of spatial resolution. All the data has been gathered from official access points, as detailed in the Data Records section. We offer provenance records that track the origin of the data and information regarding all the processing steps in those cases where the data has been consolidated. All data records are regularly-updated and are accessible through an Application Programming Interface (API) as well as through a dedicated GitHub repository. We also provide an interactive web application for exploring the data. In this work, we present the system and illustrative examples to show how the combination of mobility and health data can help public health directives to effectively mitigate COVID-19 transmission.

**Figure 1.**
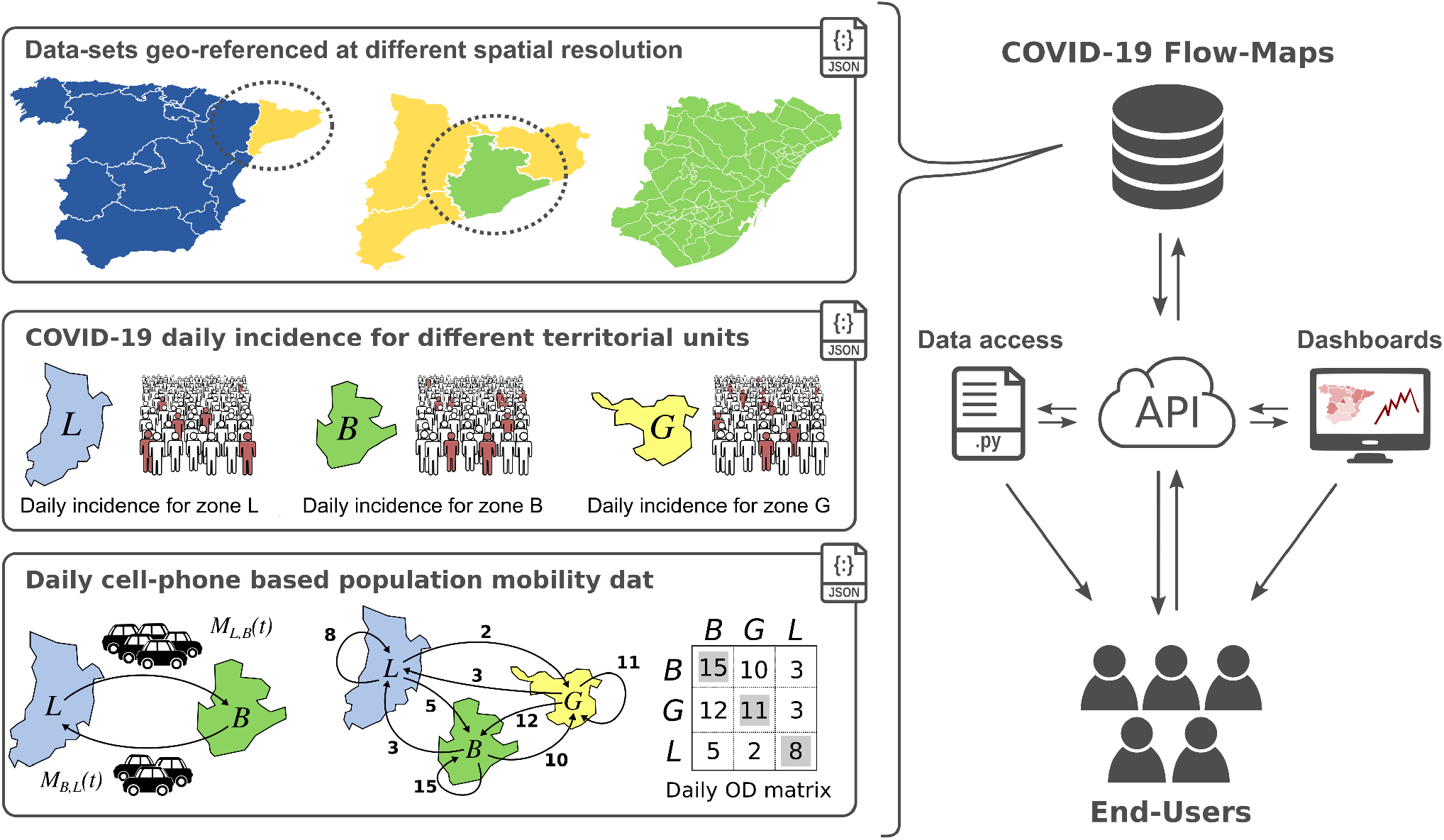
Graphical representation of COVID-19 Flow-Maps Geographic Information Systems. The main data records include geographical layers for different territorial units, COVID-19 daily cases reported at different spatial resolution and phone-based anonymized mobility data in the form of daily origin-destination matrices. All the information is stored in non-SQL database that can be directly queried through a REST-API, downloaded using provided scripts, and accessed through web-based interactive data dashboards.

## Methods

### Data acquisition

The COVID-19 data records for Spain are retrieved from endpoints provided by a variety of official sources including national-level reports, as well as those reported by autonomous community governments (see COVID-19 data and Data Records section and Online Table 1). As data is obtained from different sources, each record differs in its format, including different fields, field names, and the level of aggregation (e.g. age, gender). Additionally, data retrieved from different sources are mapped into different geographical layers (e.g. autonomous communities, provinces, municipalities). Therefore, in addition to the COVID-19 reported cases, the spatial information needed for its mapping is also retrieved from the corresponding official sources (see Online Table 2). Population mobility records based on AMPD are reported by the Spanish Ministry of Transport, Mobility and Urban Agenda (MITMA, Ministerio de Transportes, Movilidad y Agenda Urbana) in tabular format and it include two different daily reports, as well as the geographical layer needed to map the data (for further details see Mobility data in Data Records section).

**Table 1.** Table describing the different geographical layer on which main data records are reported

| Layer name | ID | Category | Polygons |
| --- | --- | --- | --- |
| Autonomous communities | cnig_ccaa | Administrative | 19 |
| Provinces | cnig_provincias | Administrative | 52 |
| Municipies | cnig_municipios | Administrative | 8212 |
| Districts | ine_districts | Administrative | 10483 |
| Cataluña Basic Health Areas | abs_09 | Sanitary | 374 |
| Madrid Basic Health Areas | zon_bas_13 | Sanitary | 286 |
| Castilla y León Basic Health Areas | zbs_07 | Sanitary | 247 |
| Navarra Basic Health Areas | zbs_15 | Sanitary | 57 |
| País Vasco Health Areas | oe_16 | Sanitary | 135 |
| Mobility Areas | mitma_mov | Custom | 2850 |

**Table 2.** Table describing the different sources of COVID-19 cases and the layer in which the data is reported. Many sources provide more information besides the incidence.

| Region | Layer | ID | First record | Extra info. |
| --- | --- | --- | --- | --- |
| Spain | Autonomous communities | ES.covid_cca | 2020-01-01 | ✓ |
| Spain | Provinces | ES.covid_cpro | 2020-01-01 | ✓ |
| Cataluña | BHA | 09.covid_abs | 2020-01-20 | ✓ |
| Navarra | BHA | 15.covid_abs | 2020-03-25 |  |
| Castilla y León | BHA | 07.covid_abs | 2020-02-29 | ✓ |
| País Vasco | BHA | 16.covid_abs | 2020-03-21 |  |
| Madrid | BHA | 13.covid_abs | 2020-02-27 |  |
| Comunidad Valenciana | Municipalities | 10.covid_cumun | 2020-05-27 | ✓ |
| Cantabria | Municipalities | 06.covid_cumun | 2020-03-30 | ✓ |
| Asturias | Municipalities | 03.covid_cumun | 2020-02-29 | ✓ |

### Data Processing and Consolidation

To automatically update the COVID-19 Flow-Maps data records we have implemented a workflow that retrieves data from different endpoints, processes and stores new records (see Code availability section). The first step in this workflow is to validate new entries by checking inconsistent and missing values. Secondly, the system controls for duplicated entries ensuring that for any given territorial unit on a given date a single reported value exists. Finally, for all the integrated data sources, the original entries, as well as the registry of any modification, are also stored in the provenance collection (see Data Provenance subsections). For each geographical layer needed to geo-localise each data entry we stored all the geometries, translating the Geographic Coordinates on ETRS89 Datum (EPSG:4258), and assigning them a symbolic and unique layer name (see Geographical Layers in Data Records section).

### COVID-19 cases reports

When integrating a source that reports COVID-19 cases, three key attributes are first identified, namely, the date for the reported event, the reported value (e.g. daily/accumulated incidence) and any attribute that can be used to geo-reference the data. Geo-referencing attributes can be geographic coordinates or, more usually, an identifier that matches a geometry of a defined geographical layer, e.g. the unique identifier of a municipality and a reference to that layer. For instance, the government of some autonomous communities reports COVID-19 cases on a daily basis at the level of Basic Health Areas (BHA) whereas others report cases by municipalities (see COVID-19 data and Data Records section). Missing values of the daily incidence for a given date or geographic area are imputed with zero values while missing values of cumulative incidence are imputed based on the previous available value. After validation, since some data sources either report daily incidence or cumulative incidence, we also calculate one attribute from the other. Moreover, field names are normalised while also keeping the original field names and values to be queried. Additionally, commonly used metrics such as accumulated incidences over one and two weeks are also calculated and stored.

### Reconstruction of mobility networks

The reconstruction of mobility networks relies on two main sources of information, namely: recorded events data and mobile phone network topology data. The former corresponds to anonymized data associated with the connection records of the mobile devices with the mobile phone network. These records include both active and passive events. Active events are made up of CDRs (Call Detail Records) that provide a record every time a device interacts with the network (calls, sending text messages, data sessions). These records are joined with passive event data (periodic update of the device position, changes in coverage areas, etc.), providing a very high temporal granularity. Regarding spatial resolution, location information is available at the level of the coverage area of each antenna - which implies a spatial resolution of tens or hundreds of meters in the city and up to several kilometres in rural areas, which provides an idea of the uncertainty that is introduced in the determination of the position according to the areas analysed.

The recorded events data are processed in a secure environment within the mobile operator’s infrastructure where this data is aggregated and anonymized in compliance with the existing European and Spanish legislation, e.g. LOPD-ODD (Ley de Protección de Datos). The phone network data includes the location of the communication towers which are used to obtain a Voronoi Diagram of the cellular coverage map. Additional sources used include land use information from the Spanish Land Use Information System (SIOSE), population data (i.e. the register of inhabitants by districts) and the Spanish transportation network (e.g. airports location, rail network, etc.).

The first step in the data processing workflow consists of the extraction and pseudonymisation of the mobile (cell) phone records. The pseudonymization of the records is based on the use of a one-way hash function, that is, a function that allows the calculation of an anonymized identifier from the original identifier (usually the International Mobile Subscriber Identity) in such a way that it is impossible to carry out the reverse process. Furthermore, a perfect hash function is used to avoid collisions, i.e. that two different original identifiers produce the same anonymized identifier. The anonymized phone records are stored in a secure environment within the infrastructure of the mobile operator, where the algorithm used to aggregate the anonymized data will also run.

The processing and analysis of the raw data can be divided into the following steps^21^: (i) pre-processing and cleaning of the data; (ii) construction of the sample; (iii) identification of the place of habitual residence and the place of overnight stay; (iv) extraction and inferences of activities and trips; (v) extrapolation of sample results; and (vi) generation of indicators. First of all, a pre-processing of the mobile phone data is carried out to facilitate its management, ordering and grouping the records in the most convenient way for further analysis. A data integrity check is also carried out to eliminate possible errors in the mobile operator’s data. The process of data cleaning and debugging of errors is essential to ensure the quality of the data, preventing possible source of errors from distorting the results obtained with the algorithms used for the extraction of activity and mobility patterns. To construct the sample, a selection of valid users is first made to provide information on their movements. This selection is made according to different criteria related to their phone activity so that this is sufficient to establish their behaviour patterns with an adequate level of reliability.

To identify the area of most usual home areas of the users, an analysis of the behavioural patterns is conducted over several weeks. In a similar way, the overnight stay area is also identified. This information will be used later in the sample extrapolation process. To identify activities and trips, a combination of criteria based on the duration of stay times, travel itineraries and patterns of behaviour throughout the study period, filtering intermediate stops subordinate to the journey and made between stages of the journey (e.g. intermediate stops for a transfer at a bus/train station). The result of this step is the sequence of activities and trips made on the analysed days. The information associated with each activity includes the location, the start time and the end time. As associated information, each trip includes origin (location of the activity immediately prior to the trip), destination (location of the activity immediately after the trip), the start time of the trip (time of the end of the previous activity) and end time (start time of the next activity), distance, home place, activity in the origin and activity in the destination.

The extrapolation of the sample is carried out by taking as the sampling reference the population residing in the country, according to the data from the Population Register provided by the National Institute of Statistics. Standard sample extrapolation procedures are used (similar to those used, for example, in a household mobility survey), applying expansion factors by area of residence based on the sample/population ratio for each age and gender segment in each district. Finally, the information obtained in the previous steps is presented with the required spatial and temporal resolution and the desired segmentation to generate the origin-destination matrices and the rest of the mobility indicators previously described. The generation of the mobility indicators has been carried out using specialised software developed for this purpose and all these processes are carried out within the mobile operator’s infrastructure, so that the information generated and delivered to our source, the MITMA official site for data release, is already aggregated and anonymized.

The output obtained from the processing steps is then used for generating two mobility indicators: the hourly-based OD matrices referred to as the Maestra 1, and the daily population mobility descriptions referred as the Maestra 2 (see Mobility data in the Data Records section). Both indicators are geo-referenced to a custom layer referred to as the MITMA mobility layer (see Geographical Layers in the Data Records section). Further details on the analysis and processing of mobility data are provided on the official page of the study (https://www.mitma.gob.es/ministerio/covid-19/evolucion-movilidad-big-data).

Mobility indicators are further processed to obtain more aggregated mobility metrics. Hourly-based OD matrices (Maestra 1) are aggregated to obtain daily mobility matrices. Furthermore, daily population mobility indicators (Maestra 2) that provide the number of people that have done zero, one, two, and more than two trips, are aggregated within each mobility area in a given date to estimate the total population of each mobility area on a daily basis. The total population computed in this way is stored to be used in the calculation of other descriptors (e.g. daily incidence by total population). The population inferred from Maestra 2 is estimated on a daily basis and thus it captures the fluctuations in the population due to net mobility over the year (e.g. mobility during summer vacations).

### Data Provenance

The database has all the information needed to identify the origin of the data, all the processing carried out, the original files retrieved, and the timestamp of the last update. Furthermore, copies of all the data obtained from the different sources are kept in their original formats and their source URL (if available). All this information can also be queried through the REST-API (see REST-API in the Usage Notes section).

### Data projection using geographical layers overlay

In order to combine COVID-19 daily incidence and population mobility data, both data records should be projected onto the same geographical layer. In some cases, one Polygons in one layer must be covered by a single polygon from another layer, with an exclusive overlap (e.g. municipalities are included in provinces). In other cases, the overlap between the two layers is not exclusive, which means that polygons in one layer can be covered by more than one polygon from another layer. For instance, COVID-19 daily incidences and mobility data are geo-referenced into different geographical layers that cannot be combined directly. Thus, to overcome this issue, we have implemented a general approach to project data among different geographical layers. The approach is based on linear interpolation over the overlaying areas between the polygons from the two layers. We call this process “projecting” data from a source layer (e.g. municipalities) into the target layer (e.g. BHAs).

#### Spatial Overlay Matrix

In general, a geographical layer is composed of several different polygons. For example, in the case of the province layer, each individual province will correspond to a polygon that defines its geographic frontier or border. Therefore, given two layers *A* = {*A*_1_, *A*_2_, …} with *N*^*A*^ polygons and {*B* = *B*_1_, *B*_2_, …} with *N*^*B*^ polygons, we define the *N*^*A*^*xN*^*B*^ Spatial Overlay Matrix *O*^*AB*^ where 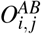 contains the proportion of the polygon *A*_*i*_ that falls intothe polygon *B*_*j*_. In general, *O*^*AB*^ ≠ *O*^*BA*^ and all the rows in the overlay matrix sum up to 1:

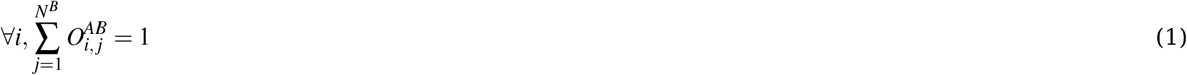

#### Population-based overlay matrix

Using Spatial-based overlays has the implicit assumption that the population is distributed uniformly on the territory. Nonetheless, this assumption may not hold in most cases, and therefore, some authors have proposed using population density grids to improve the projection’s accuracy. The idea is to account for how the population is distributed among the territory and use the population overlays, i.e. the number of people each area of the first layer share with each area of the second layer. Therefore this approach requires information on how the population is distributed with high spatial resolution. Herein we use the population density grid provided by GEOSTAT^22^ that accounts for the population distribution of all Europe. This grid has cells of 1kmš, and each cell has assigned an estimated population value.

To calculate the population-based overlays between layer A and a second layer B using the population density grid G we first calculate the intersections between A and B obtaining a set of intersections *I*_*AB*_ (as in the case of the spatial overlay). Then, for each intersection, we calculate its population by performing a second intersection with the population grid. The population of each intersection is calculated as the sum of the population of the cells that fit completely in the intersection, and the fraction of those cells that overlay but do not fit completely. Once we have the population of each intersection, we build the overlay matrix using population values instead of areas. Finally, as in the case of the Spatial-based overlays, the rows of the overlay matrix are normalised so that each row sums one (see Equation 1).

#### Data projection using the overlay matrix

Given a row vector *V*^*A*^ of data on layer A (e.g. COVID-19 incidence value for each polygon of layer A), this data can be projected into the layer B by just multiplying *V*^*A*^ and the overlay matrix:

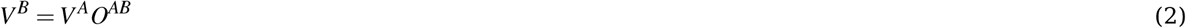

Figure 2-a shows an example of how cases geo-referenced in an origin layer **A** can be projected into a destination layer **B** using a pre-calculated overlay matrix. Interestingly, this approach does not depend on how the overlay matrix was constructed, i.e. if it is based on geographic or population-based overlays.

**Figure 2.**
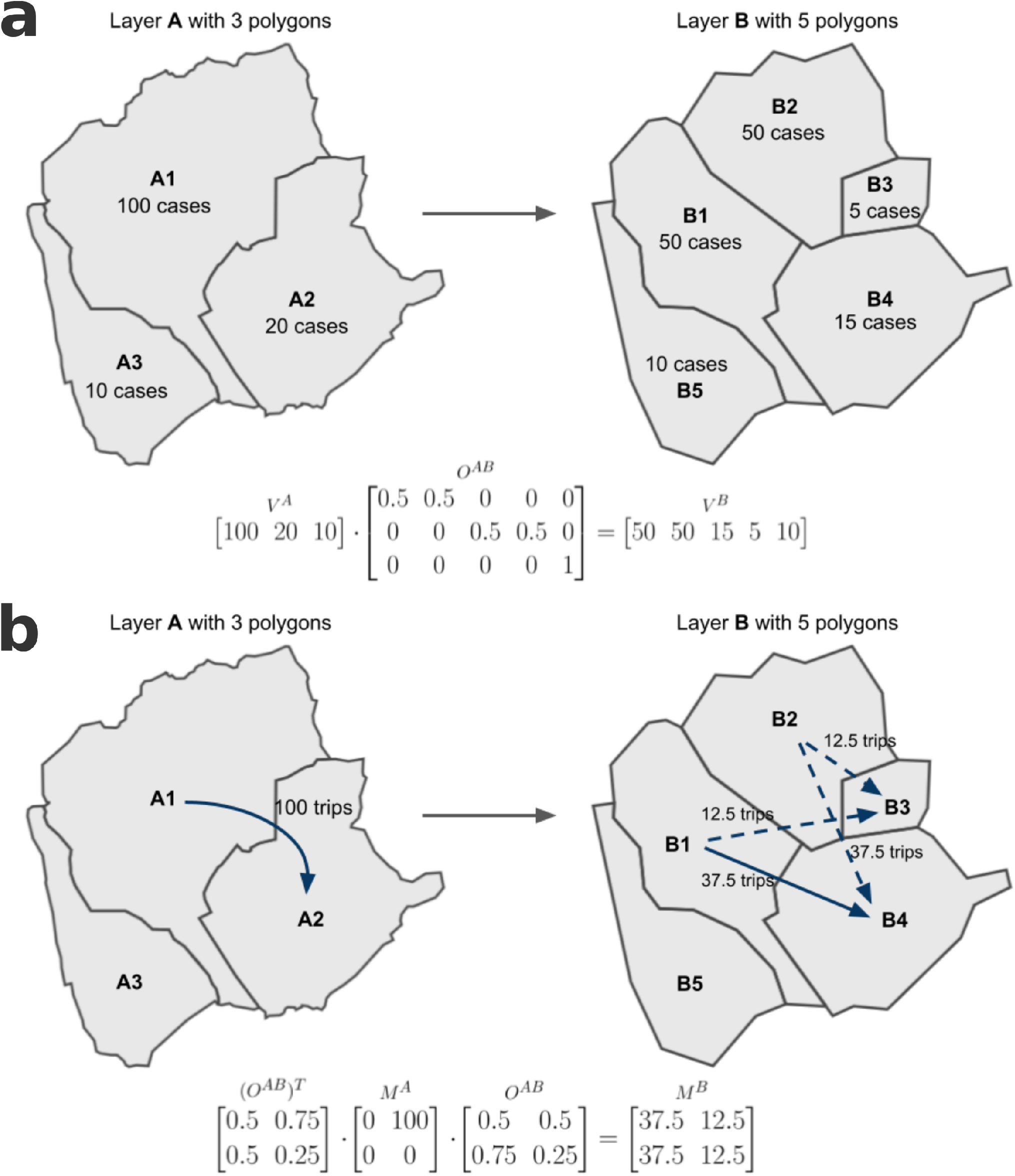
Toy example to explain the approach for projecting data between layers using Spatial-based overlays. Panel a shows an example of cases projection from layer A to layer B, using the Spatial-based overlays between both layers. Panel b shows an example of trips projection between the same layers.

The same approach is also used to project an OD mobility matrix between geographical layers. Projecting OD matrices to other geographical layer allow the integration of population mobility with data-set reported in a different layer, e.g. one on which COVID-19 cases are reported. In general, given a mobility matrix *M*^*A*^ geo-referenced to the layer A, each entry 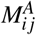 contains the number of trips from polygon *A*_*i*_ to polygon *A*_*j*_, (both polygons from layer A) it can be projected to the layer B by multiplying it by the overlay matrix and its transpose:

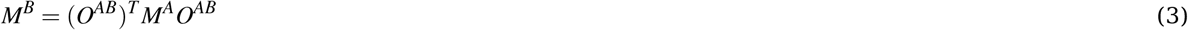

Such calculation can be seen as first projecting the origins and then projecting the destinations (2-b). Overlay matrices for all the combinations of geographical layers have been computed and stored in the database, enabling a fast projection of any data-set between the different geographical layers.

### Mobility Associated Risk

To assess the effect of population mobility on the spreading of COVID-19, we have developed a risk score named Mobility Associated Risk (MAR). The MAR score integrates daily cases with populations flows between different geographical areas (i.e. OD matrices), e.g. provinces or BHAs, to estimate how likely it is that cases spread as a consequence of human mobility (see Figure 3). Herein, we use the incidence accumulated over two weeks as an estimator of the number of active cases. Then, for a given geographical layer *L* with *n* zones *j* := 1…*n*, and a given date *t*, we refer to the cases accumulated over two weeks as 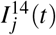 This estimator is then normalised to the total population reported in zone *j* at the same date *t*.

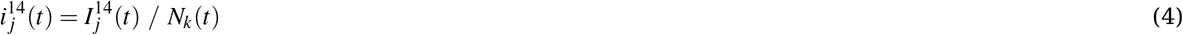

where *N*_*k*_ (*t*) is the total population in zone *j* at date *t* and *i* _*j*_ (*t*) is the estimator of the active cases per total number of inhabitants. We then combine the *i* _*j*_ (*t*) from each zone *j* together with a daily OD mobility matrix for the date *t* as follows:

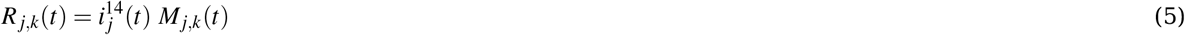

where *M*_*j,k*_ (*t*) is the number of trips from *j* to *k* with both values reported at date *t*, and *R*_*j,k*_ (*t*) is an estimation of the expected number of infected subjects also moving from zone *j* to *k* at day *t*. In general, when the daily COVID-19 cases and the mobility are reported in the same geographical layer *L*, the risk *R*_*j,k*_ (*t*) can be calculated for all pairs of zones *j* and *k* by the element-wise, or Hadamard product between the *nx*1 vector 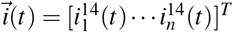 of cases densities and the *nxn* mobility matrix *M*(*t*):

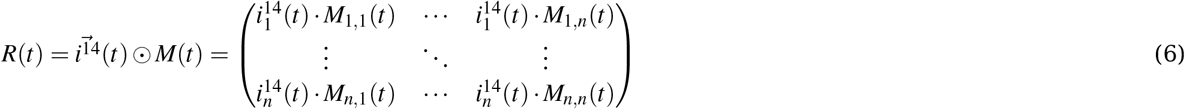

**Figure 3.**
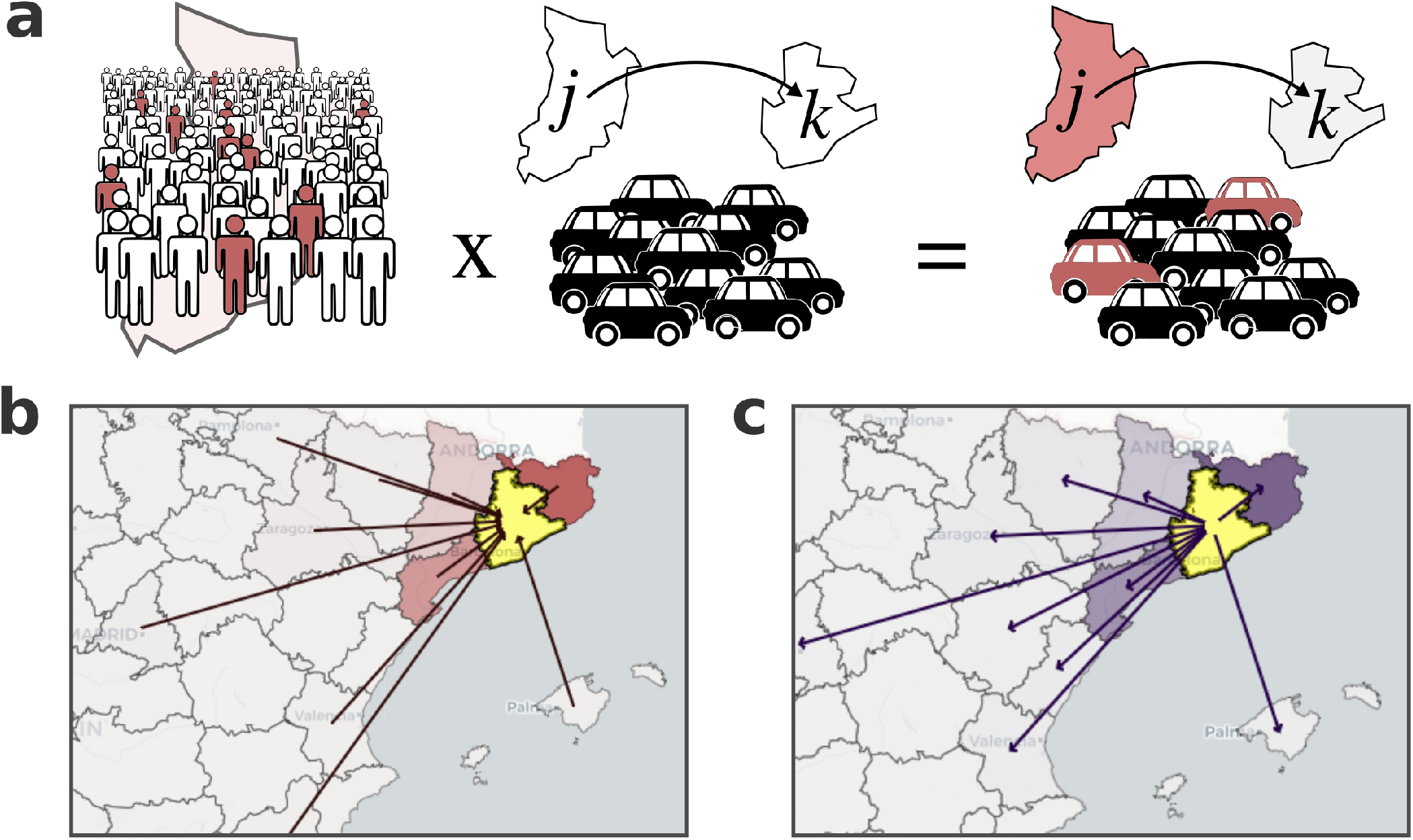
Mobility Associated Risk chart. Panel a is a graphic representation of how the mobility associated risk between zones *j* and *k* is calculated from the normalised accumulated incidence and the number of trips. Panels b and c show nd example of the total incoming and outgoing risk for the province of Barcelona (highlighted in yellow). The colour scale on the maps from (c) and (b) indicate the incoming MAR and outgoing MAR, respectively; more intense red/violet indicate a greater incoming/outgoing MAR respect the target zone. Arrows indicate the top ten zones of incoming (b) outgoing (c) MAR.

The matrix *R*(*t*) is thus a directed weighted network where the nodes correspond to the different *n* zones from layer *L* and the flow *R*_*j,k*_ (*t*) between any pair of nodes (*j, k*) is the estimated number of infected subjects moving from the source *j* to target *k* at *t*. The network-based structure of the mobility associated risk allows the definition of the total MAR incoming to zone *k* by summing the risk over all possible sources (i.e. summing the *kth* column of *R*(*t*)). In a similar way, that outgoing MAR for a given zone *k* corresponds to all the weighted edges having *j* as the source node (i.e. summing the *kth* row of *R*(*t*)).

The risk network can be represented in the map to analyse the source of incoming MAR to any zone of interest. For instance, the risk network *R*(*t*) can be calculated between provinces by combining daily incidence reported at province level together with mobility data aggregated at the same level. Figure 3b-c shows a representation of the incoming and outgoing mobility-associated risk for the province of Barcelona for the 10th of October 2020. The incoming risk represents the expected number of imported cases from other provinces (Figure 3-b) whereas the outgoing risk corresponds to the expected number of infected individuals travelling to a different province (Figure 3-c). Mobility Associated Risk (MAR) can be calculated at different scales of spatial resolution by aggregating mobility zones based on different criteria.

Finally, we would like to stress that although we are aware that the MAR score is a raw approximation (e.g. infected cases in quarantine are not expected to travel with the same frequency as healthy people does), we find it can be used as an approach to evaluate the risk of outbreaks in different zones due to imported cases.

### Data Visualisation

We have developed a web interactive data dashboard to visualise different metrics as well as results from different analysis pipelines using interactive maps, plots and tables. The COVID-19 Flow-Maps Boards are a set of interactive web dashboards that provide access to different reports of the health situation, the population mobility and its associated risk for the different regions of Spain. Currently, we provide access to three different interactive dashboards for the analysis and visualisation of the evolution of COVID-19 cases in Spain (at three different scales of spatial resolution); the population mobility between different municipalities (or districts, in densely populated areas); and the MAR networks between regions. The COVID-19 Flow-Maps Boards can be accessed through the following link: http://flowmaps.life.bsc.es/.

## Data Records

COVID-19 Flow-Maps (http://flowmaps.life.bsc.es/) is a geographic information system that integrates two main sources of information for analysing the COVID-19 pandemic in Spain. In the first place, the system provides daily reports on COVID-19 cases for different regions and at different scales of spatial resolution; secondly, the system provides access to a daily updated data-set on population mobility which includes Origin-Destination (OD) matrices and the total number of trips per person per day. The system also provides access to the different geographical layers to which the different data-sets are geo-referenced. All data collections are stored in a distributed MongoDB database that can be queried through a REST-API and a command-line utility (see Usage Notes section). Furthermore, the data records introduced in this section can be downloaded from our periodically updated GitHub data repository (https://github.com/bsc-flowmaps).

### Geographical Layers

This data record includes all geographical layers on which the different data records are geo-referenced (e.g. mobility, COVID-19 cases). The different layers can be grouped into those that cover the whole territory of Spain (e.g. municipalities) and those that are restricted to a specific region (Table 1). Among those that cover the full territory of Spain, the record accounts for the first four levels of administrative division, that is, autonomous communities, provinces, municipalities and districts. These layers are retrieved from the National Centre of Geographic Information (see Online Table 2 for details on the sources). Additionally, in the case of the mobility data-set, the events are geo-referenced to mobility zones from a custom layer that is provided together with the mobility data-sets. This layer was reconstructed combining cell-phone antennas coverage areas together with districts and municipalities and includes 2850 different mobility areas (see Figure 4-c) that cover almost the entire territory of Spain. In general, each area in the MITMA mobility layer may correspond to a district or group of districts in densely populated areas, and to municipalities or groups of municipalities in regions with low population density. However, there are some rural areas not covered by cell phone antennas and thus not assigned to any mobility zone.

**Figure 4.**
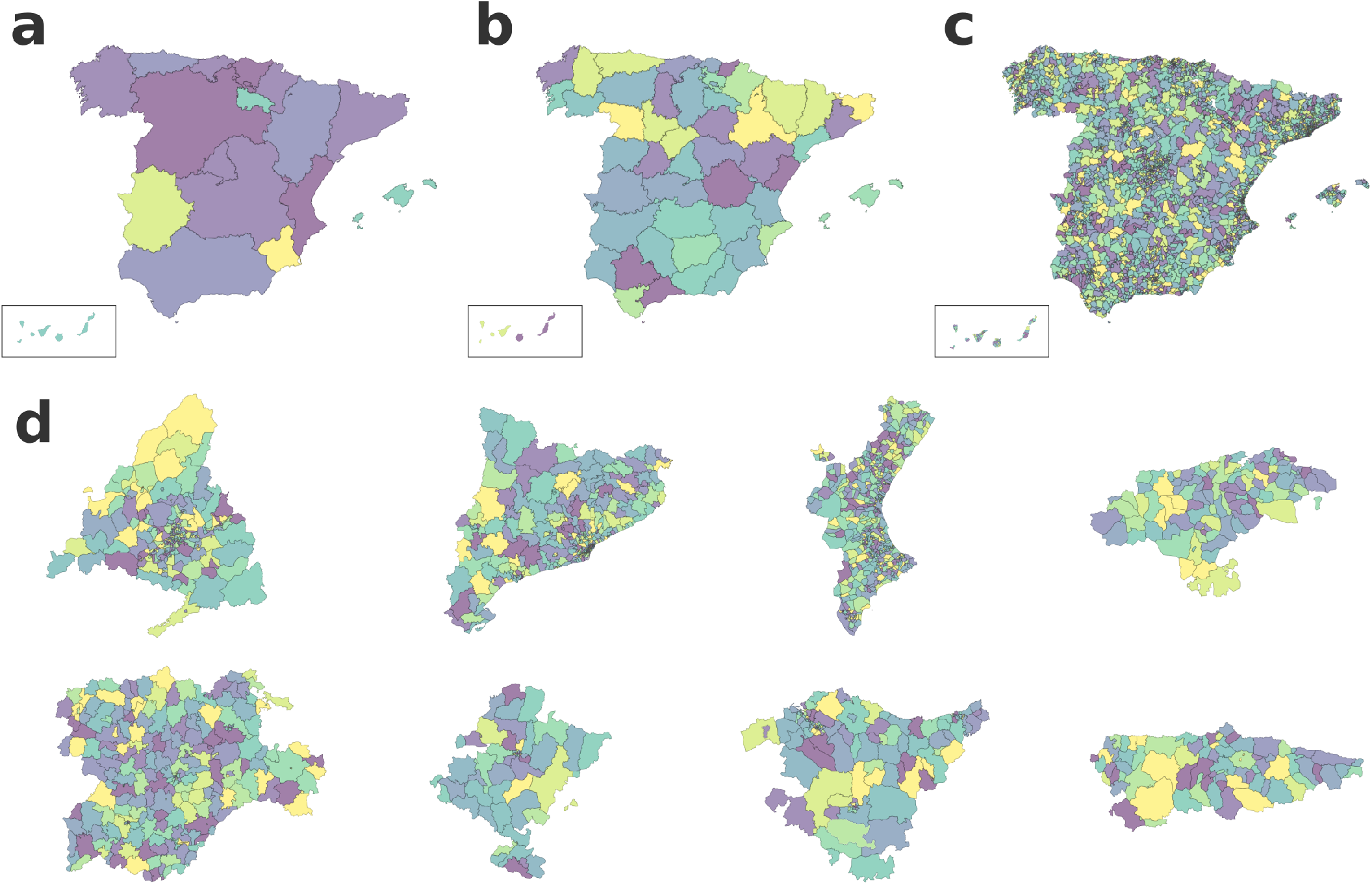
The figure represents the different geographical layer included in the database. In (a) and (b) the coloured polygons correspond to the different autonomous communities and provinces of Spain, respectively. For simplicity, the Canarias islands are represented in the bottom left box. Panel (c) represents the MITMA mobility layer and the coloured polygons correspond to individual mobility zones that match district in high-density populated areas and municipalities or groups of municipalities in less populated areas. Panel (d) represents the layers for which some autonomous communities report COVID-19 cases with higher spatial resolution than province level. From top to bottom and left to right the layers: Madrid’s BHAs, Cataluña’s BHAs, Valencia’s Municipalities, Cantabria’s municipalities, Castilla y León BHAs, Navarra’s BHAs, País Vasco BHAs, Asturias’ municipalities. In all the plots colours are only used for visualisation purposes.

The set of layers for specific regions corresponds to the Basic Health Areas (BHA) of different autonomous communities (see Figure 4 d). According to the national health administration of Spain, each autonomous community is divided in a set of BHAs, where each individual area corresponds to the geographic delimitation for the operating of a primary care unit. Because many autonomous communities report COVID-19 cases by BHA, we have included the corresponding layer. Table 1 only includes the geographical layer of the BHA for Cataluña, Castilla y León, Madrid, Navarra and País Vasco, because those are the regions that report COVID-19 cases at the levels of BHAs. The data record also includes other geographical layers (see COVID-19 data in the Data Records section). Geographical layers are distributed in GeoJSON format in one layer per file. In addition to the coordinates of polygons, each layer file also provides the information needed for it crosse-reference with the other data-records, including unique polygon identifiers, province and autonomous communities unique codes, as well as other useful information such as pre-calculated centroids. The released data can be retrieved using the following link: https://doi.org/10.5281/zenodo.4634663.

### COVID-19 data

The COVID-19 record includes daily cases for all Spain reported at the level of autonomous communities as well as provinces (see Figure 4 a and b). Each record has an associated date, the corresponding identifier of the layer and code of the region and a set of COVID-19 related fields, which include the number of cases (daily incidence) as well as the number of cases segregated by test type, i.e. PCR, antigen, antibody, ELISA or unknown. Additionally, information on the total daily hospitalisations and admissions into intensive healthcare units, reported by provinces, are also provided.

This data record also includes COVID-19 cases at a higher spatial resolution (e.g. municipalities) reported by several autonomous communities. Currently, eight out of the nineteen autonomous communities publish reports with local daily COVID-19 cases at the level of municipalities or BHAs. On the one hand, Castilla y León, Madrid, Navarra and País Vasco report COVID-19 cases at the level of local BHAs; on the other hand Asturias, Navarra and Valencia report daily access at the level of municipalities; Cataluña local government reports COVID-19 daily cases at the level of BHA as well as municipalities. Table 2 shows the different COVID-19 data-sets that are integrated into this record, together with the associated geographical layer in which the data is reported. In this way, each entry reporting COVID-19 cases includes the reporting date, the geographical layer and identifier of the specific polygon within the layer and the number of cases reported, reported as daily and cumulative incidence. Additionally, each entry also includes useful metrics, such as the rolling average of the daily incidence over one and two weeks, the population in that area and the number of cases per 100,000 inhabitants. More detailed information regarding the specific data fields reported by each different source is provided in Online Table 3. The released data can be retrieved using the following link: https://doi.org/10.5281/zenodo.4634869.

**Table 3.** Table describing the different layer for which OD matrices have been projected into.

| data-set ID | Origin layer | Target layer | Projection Type | Frequency |
| --- | --- | --- | --- | --- |
| mitma_mov_mitma_mov | mitma_mov | mitma_mov | None | Hourly |
| mitma_mov_mitma_mov | mitma_mov | mitma_mov | None | Daily |
| abs_09_abs_09 | abs_09 | abs_09 | Population-based | Daily |
| abs_09_cnig_provincias | abs_09 | cnig_provincias | Population-based | Daily |
| cnig_ccaa_cnig_ccaa | cnig_ccaa | cnig_ccaa | Population-based | Daily |
| cnig_provincias_abs_09 | cnig_provincias | abs_09 | Population-based | Daily |
| cnig_provincias_cnig_provincias | cnig_provincias | cnig_provincias | Population-based | Daily |
| cnig_provincias_oe_16 | cnig_provincias | oe_16 | Population-based | Daily |
| cnig_provincias_zbs_07 | cnig_provincias | zbs_07 | Population-based | Daily |
| cnig_provincias_zbs_15 | cnig_provincias | zbs_15 | Population-based | Daily |
| cnig_provincias_zon_bas_13 | cnig_provincias | zon_bas_13 | Population-based | Daily |
| oe_16_cnig_provincias | oe_16 | cnig_provincias | Population-based | Daily |
| oe_16_oe_16 | oe_16 | oe_16 | Population-based | Daily |
| zbs_07_cnig_provincias | zbs_07 | cnig_provincias | Population-based | Daily |
| zbs_07_zbs_07 | zbs_07 | zbs_07 | Population-based | Daily |
| zbs_15_cnig_provincias | zbs_15 | cnig_provincias | Population-based | Daily |
| zbs_15_zbs_15 | zbs_15 | zbs_15 | Population-based | Daily |
| zon_bas_13_cnig_provincias | zon_bas_13 | cnig_provincias | Population-based | Daily |
| zon_bas_13_zon_bas_13 | zon_bas_13 | zon_bas_13 | Population-based | Daily |

### Mobility data

Mobility data records come from a study conducted by MITMA, which analyses the mobility and distribution of the population in Spain from February 14^th^ 2020 until the present date, and will be conducted until December 31^st^ 2021. (https://www.mitma.gob.es/ministerio/covid-19/evolucion-movilidad-big-data). The study is based on a sample of more than 13 million anonymised mobile-phone lines provided by a single mobile operator whose subscribers are evenly distributed.

The data record includes two different and complementary indicators, referred to as Maestra 1 and Maestra 2, whose records are geo-referenced to the MITMA mobility layer (see Table 1). The MITMA layer is composed of 2850 zones and is described in Geographical Layers within the Data Records section. The unit to measure mobility is the trip. For any person, it is considered as a trip any movement of more than 500 meters that lasts more than 20 minutes (see Data Processing and Consolidation). Is worth noting that, the origin and destination zone might be the same one (e.g. a person that goes to the store nearby his/her residence).

#### Maestra 1: Origin-Destination matrix for the mobility layer, with hourly resolution

Each entry has a date and time period (the range between two consecutive hours), the origin and destination zones and the number of trips from origin to destination. Origin and destination zones correspond to geometries from the MITMA mobility layer and internal trips (same layer of origin and destination) are also reported. Moreover, each entry also reports the putative activity of the grouped trips (from home to work, for instance), ranked travelled distances (in kilometres) as well as the total distance covered by the aggregated trips. The activity at the origin and at the destination for the reported trips in a given entry is classified into one of the following options: home, work, or other; therefore, each data entry groups trips by origin-destination and by all the matrix segmentation (place of residence, distance, travel time, activity at the origin, activity at the destination, etc).

#### Maestra 2: Trips per person matrix on each mobility area on a daily basis

This indicator reports population-based daily mobility behaviour. For each date and zone from the MITMA mobility layer, the indicator reports how many persons have performed 0, 1, 2 or more than 2 trips. While the indicator does not provide the destination of the trips, it accounts for the fractions of people performing at least one trip or none, as well as the estimated total population in that zone for the given date (considering as population those persons who stay overnight in the zone on that date).

OD matrices have been projected into different geographical layers and at different spatial resolution and are provided to direct use. Table 3 describes the different origin/destination layers into which OD matrices have been projected and are provided. The released data can be retrieved using the following link: https://doi.org/10.5281/zenodo.4634896. Mobility data with hourly resolution can be retrieved from the COVID-19 Flow-Maps system using the REST-API. Regardin population data, the released data can be retrieved using the following link: https://10.5281/zenodo.4635259

## Technical Validation

### Validation of estimated population

Population values used for different calculations (e.g. daily cases per total population, mobility associated risk) are estimated using the indicator Maestra 2 from the mobility data record where the population in each MITMA mobility zones is daily reported. We use this estimation because it accounts for the population fluctuations in the different regions over the year. Herein, we compare our estimated population values to those reported in the Spanish census of 2019^23^. The population values from the Spanish census are reported by municipalities whereas our estimations are reported by MITMA mobility areas. To allow the comparison of both data-sets, we aggregate population to province-level and compare the values at this geographical level. Moreover, we compare the population reported in the census with the population estimated from Maestra 2 indicator for four different dates. We calculate the Person correlation and found coefficients greater than 0.99 in the four cases. We also calculate the mean relative difference and in the four cases we find values lower than 2.5% (see Online Table 4). The difference may be due to the fact that some people do not live in the city where they are registered, among other things. Furthermore, the results show that the estimated values that are more similar to those reported by the census, correspond to the population estimated for the 2020-04-15, which is just after the beginning of the lockdown. We also find that population estimates that exhibit larger deviations from the reference values are for those of the autonomous cities of Ceuta and Melilla that have a very particular population structure (see Online Table 4). In summary, we find that the population estimated from Maestra 2 indicator is in good agreement with those values reported in the Spanish census of 2019.

### Evaluation of Spatial and Population-based data projection approaches

As explained in the Data projection using geographical layers overlay subsection within the Methods section, data projection is a critical step for the integration of data-sets reported in different geographical layers. The projection is a linear interpolation using a defined overlay matrix. We have implemented two different approaches to reconstruct overlay matrices; one approach uses the ratios between areas (Spatial-based) whereas the other introduces a correction based on the distribution of the population (Population-based). To compare the accuracy of both approaches we first project the population reported by MITMA mobility areas into municipalities using both, Spatial and Population-based overlay matrices. Second, we compare the population values resulting from the projection into municipalities from those of the Spanish census of 2019^23^. Figure 5 shows the comparison between the population projected using both overlay matrices with respect to those of the Spanish census. In both cases, we find correlation coefficients close to one. Nonetheless, the population projected the Spatial-based overlays exhibit a higher dispersion. To quantify these differences we calculate the RMSE between the projected values and those from the census. The RMSE found for the Spatial-based and Population-based overlays are 3200 and 2500, respectively. Thus, although Spatial-based overlays seem to be a good approximation, population-based overlays introduce less distortion and produce more reliable results.

**Figure 5.**
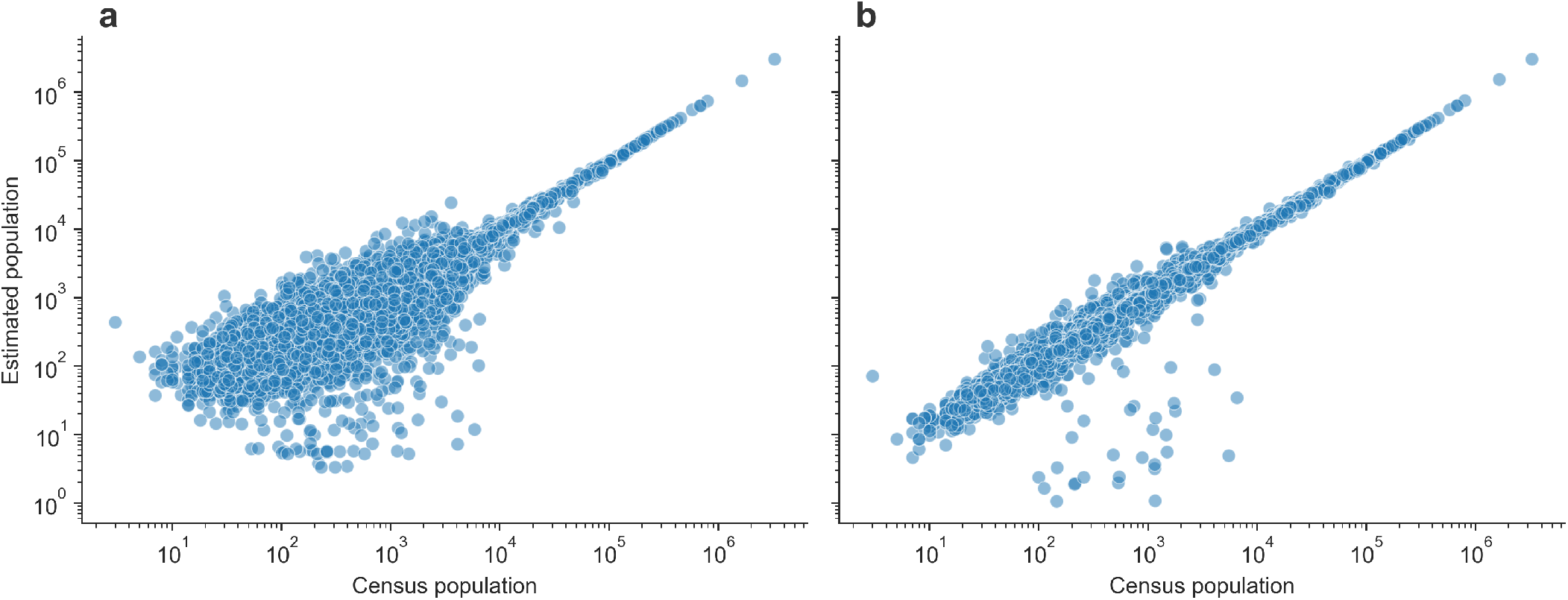
Comparison of data projection approaches between geographical layers. The figure represents the census population reported by municipalities with respect to those values estimated from MITMA mobility data after its projection from the MITMA mobility into the municipalities layer. Panel (a) and (b) show the result of the projection using based Spatial and Population-based overlays, respectively.

### Comparison of OD mobility data with other studies

To evaluate the quality of the mobility data from MITMA we compare our daily OD matrices to those from a different data-set. For this comparison, we use data from a study conducted by the National Institute of Statistics (INE, Instituto Nacional de Estadística). This study is based on the analysis of the position of more than 80% of mobile phones throughout Spain and it provides OD flows between areas and between mobility areas as long as they involve more than 15 people^24^. The data-set includes daily reported OD matrices from March to June 2020 and the geographical layer in which the population flows are reported is very similar to MITMA mobility layer. Nonetheless, to compare both data-set we first identify the common set of mobility zones and find 2680 mobility zones common to both data-set. Using the set of common mobility areas, we compare the number of trips for every origin-destination combination in the data. We also aggregate the mobility network at provinces and autonomous communities levels and compare the resulting aggregated OD matrices. Moreover, we compare both mobility data-sets for four different dates. Figure 6 shows the results of the different comparisons. These comparisons show high correlations in all cases. Nonetheless, while the comparison of the aggregated OD matrices at both, province and autonomous community levels, show a very high correlation (*R*^2^ ∼ 1), the comparison at the mobility areas level shows more dispersion (*R*^2^ ∼ 0.8). As expected, the discrepancies increase when lower fluxes are compared and decrease when higher ones are considered. Strikingly, the slope of the linear regression is ∼ 10 in all the cases, indicating the MITMA data-sets measure more trips than those reported in the INE study. This can be due to the fact that in the INE data-set the definition of trip differs from that used in the MITMA study; specifically, in the INE study, the destination areas are defined as those where the phones are more frequently detected during the 10 am to 4 pm period of a given day, as long as the phone has been in that area for at least two hours^24^. Beside these systemic differences, the comparison shows that there is a good agreement between both studies. Nonetheless MITMA mobility data record, here presented, offers trips reported on an hourly basis with a more accurate trip definition and the study, starting in February 2020 is expected to continue reporting data.

**Figure 6.**
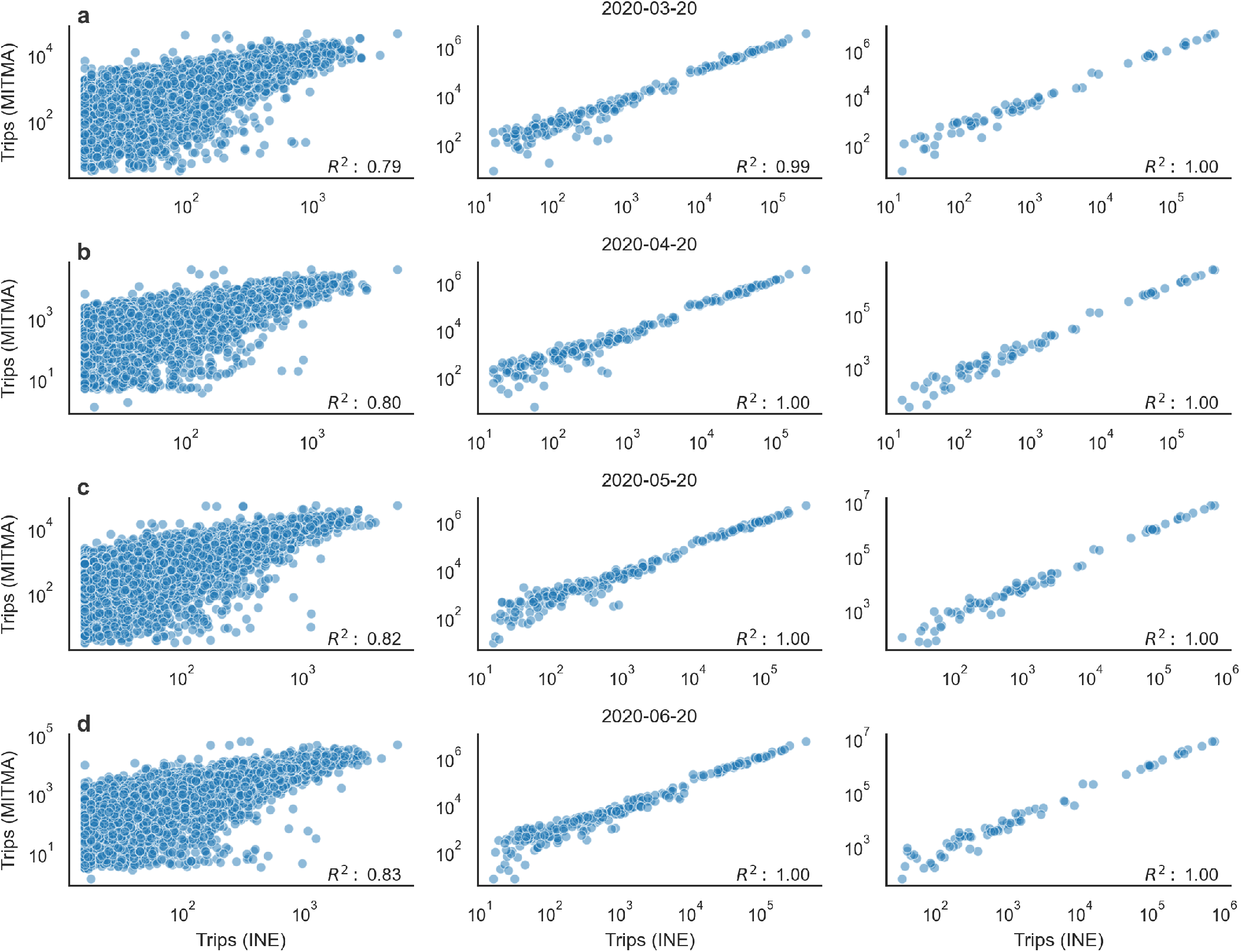
Comparison of mobility data-sets. The figure shows the comparison between the number of trips reported by the MITMA and the INE data-sets at different levels of aggregation. From left to right, each panel presents the comparison at mobility areas level, province level, and autonomous communities level. From (a) to (d) the plots represent the comparison between different dates indicated on the top of each panel. The Pearson correlation coefficient is indicated on the bottom right corner of each panel.

## Usage Notes

In this section, we present simple examples to illustrate the potential uses of this data source, as well as how to access the different data records. In addition, we also provide some suggestions on how to integrate and analyse the different data records. Data records can be accessed in three different ways. We have implemented a REST-API to allow querying the whole COVID-19 Flow-Maps system (see REST-API in Code availability section); on top the REST-API we developed command-line download toolkit to download the data records in a more simple way (see REST-API Code availability section); in addition, all data records are periodically uploaded to our data repository (https://github.com/bsc-flowmaps). Finally, we have also developed a set of web-based interactive data dashboards for exploratory analysis of the data records (https://flowmaps.life.bsc.es/flowboard/).

### COVID-19 Incidence

The COVID-19 data record includes cases reported at different levels of spatial resolution (Figure 7). Further, exploratory analysis shows that data-sets on COVID-19 cases reported by different sources may have different reporting biases. For instance, the data-sets that account for all of Spain, i.e. ES.covid_cca and ES.covid_pro do not report cases on weekends and these cases are reported on Mondays. To avoid this kind of biases we suggest users to apply rolling means on the time series previous to and analysis. Alternatively, users can also use weekly accumulated incidence for any given date. Weekly accumulated incidence can also be used as a proxy to estimate the number of infected individuals in a given date; a useful metric for epidemiological modelling. In general, we have found that the criteria used for reporting can vary between the different sources included in this data record. Furthermore, we find that in order to evaluate the health state of a particular region in absolute numbers, weekly accumulated or averaged incidence are good estimators. However, to compare the incidence between different regions we suggest the use of cases per 100,000 inhabitants to normalise by populations size. In Figure 7 we use maps to show cases reported in different regions; the maps on the left-side panel show daily incidence whereas those on the right side show cases per 100,000 inhabitants. To facilitate the analysis of the data the weekly accumulated incidence as well as cases per 100,000 inhabitants have been pre-calculated and included in all the provided COVID-19 data-sets. The COVID-19 data record of Flow-Maps also provides cases reported at higher spatial resolution. For instance, Figure 7 shows two incidences metrics at the level of provinces for all of Spain and at the level of the BHA of Cataluña. The resulting plot shows both the net incidence and the incidence by 100k population. We find maps are powerful visualisation tools and for that reason, we also provide geographical layers in GeoJSON format (see Geographical Layers in Methods sections). All COVID-19 data-set are cross-referenced to the corresponding layer in which the data is reported which facilitate the use of maps for visualisation proposes. Herein, maps were generated using Plotly library (see Code availability for further details on the tools used). To facilitate the exploratory analysis of COVID-19 data-sets our COVID-19 Flow-Board web application provides an integrated dashboard for visualising incidence plots, including interactive maps and time series curves. This analysis tool also allows the users to explore data on cases for different rates and at different scales of spatial resolution and can be accessed through the following link: https://flowmaps.life.bsc.es/flowboard/board_incidence/.

**Figure 7.**
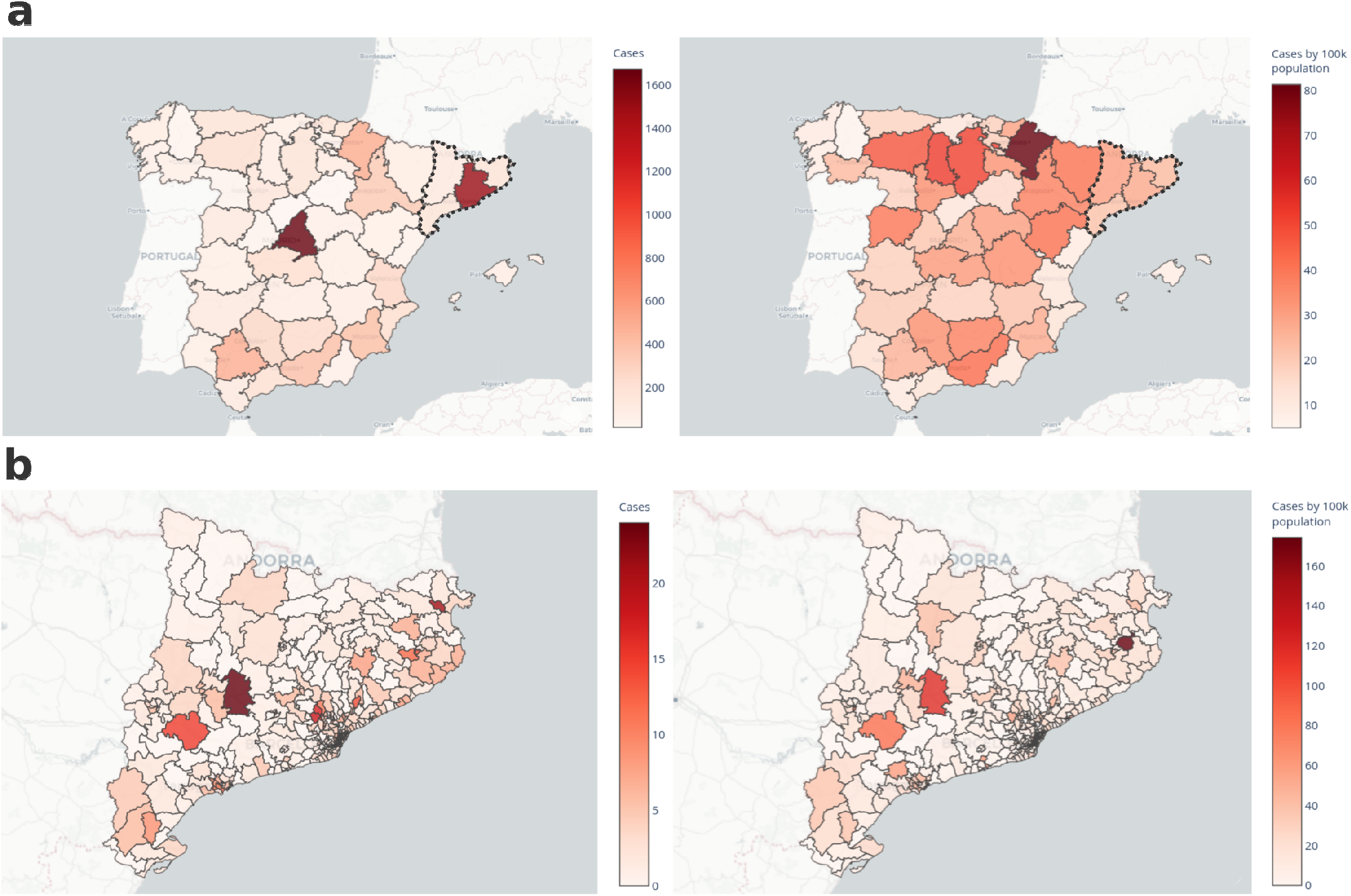
COVID-19 cases for different spatial resolutions. Panel a and b show daily incidence and accumulated incidence in a week per 100.000 people reported at the level of province and Cataluña BHA, respectively.

### Mobility data analysis

To evaluate population mobility in general terms we use as an index the percentage of people taking at least one daily trip. This mobility index is more accurate than the total number of trips because a single person can perform several trips in a given date. Figure 8-a shows the result of a coarse-grained analysis of population mobility from 14^th^ February of 2020 to 15^th^ January of 2021 in Spain, using the mobility index. The results show a seasonal behaviour with a drop on the weekends. For this reason, when comparing mobility for different dates, we suggest comparing dates that correspond to the same day of the week. Moreover, the mobility index shows an abrupt change at the begging of the lockdown that lasted from 14^th^ March until the 21^st^ Jun of 2020 (Figure 8-a. The analysis also shows that the number of people taking no trips doubled, while the number of people taking more than two trips was reduced to half (data not shown). Another change in the mobility patterns can be noted at the beginning of January when the Iberian Peninsula was hit by an extraordinary snowstorm, the largest in Spain since 1971^25^. As a consequence, population mobility was severely reduced due to the state of the roads and highways.

**Figure 8.**
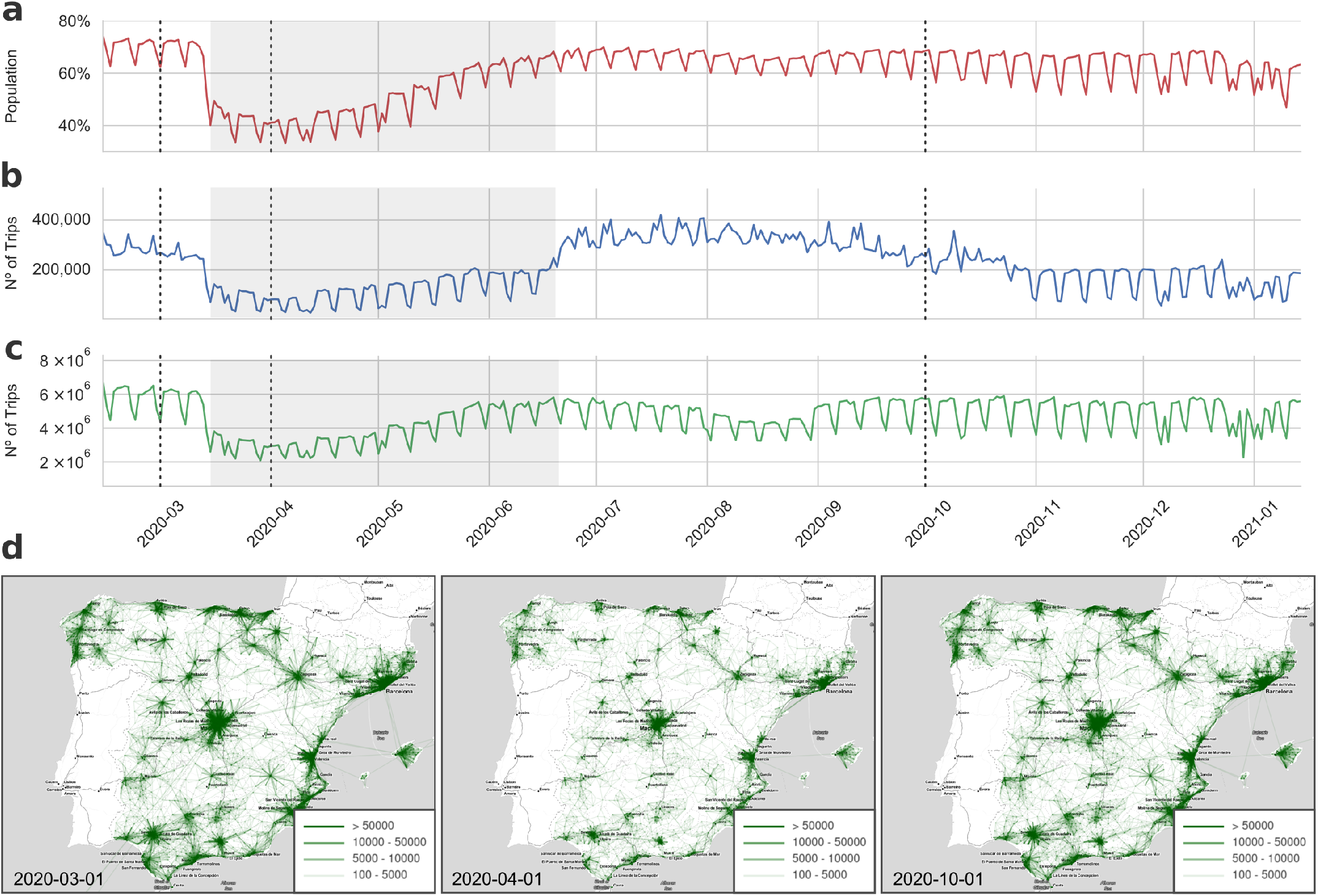
Population mobility patterns in Spain during 2020. Panel a shows the population percentage that performs one or more trips in a given day. Panel (b) and *c* show the total number of daily trips between, and within autonomous communities, respectively. Grey shaded area corresponds to the strict lockdown, whereas dashed lines indicate the date of the mobility networks represented in panel (d). Panel (d) represents the mobility networks between MITMA zones for three selected dates that are annotated on the left bottom corner and indicated with a dashed vertical line in panels (a-c).

On the other hand, OD matrices should be used when analyzing population mobility flows between different regions. Figures 8-b-c show the number of trips within and between autonomous communities. While the analysis of the flows also show a very similar pattern to the one exhibited by the mobility index (Figures 8-a), the analysis by flows also indicate change during summer. For instance, an increase in the number of trips between autonomous communities is observed in summer whereas the opposite trend is found for internal trips. Finally, to illustrate the granularity of the mobility data Figure 8-d shows a representation of the origin-destination network at the level of MITMA mobility areas for three different states: before the pandemic, during the lockdown, and after the summer. To facilitate the exploratory analysis of the mobility data-sets the COVID-19 Flow-Board provides an integrated dashboard for visualising map-based representation of the mobility networks at different scales of spatial resolution. This analysis tool basic data analytics on population mobility and can be accessed through the following link: https://flowmaps.life.bsc.es/flowboard/board_mobility/.

### Mobility Associated Risk

In order to integrate mobility and COVID-19 cases, we have introduced a risk score named Mobility Associated Risk. The MAR score combines mobility flows and incidence to estimate the potential risk of importing/exporting cases (see Mobility Associated Risk in Methods section). Thus, the MAR score could be of potential use to assist the design of targeted NPI, e.g. confining a specific region, or to inform citizens about the current situation. Moreover, the MAR score can be used to compare the different source of incoming risk for a given region. For instance, Figure 9 shows the evolution of the MAR score incoming to Asturias during 2020. We have selected the top four sources of risk. To illustrate the idea underlying the MAR score, the Figure shows the evolution of cases in each source together with the number of trips from the source to Asturias (9-a-b). Combining the former indicators the MAR score is calculated (see Figure 9-c). The Figure allows seeing that during summer, the main source of incoming MAR for Asturias was Madrid. Interestingly, for those dates, the number of trips coming from Madrid is considerably lower than the other three sources. Nonetheless, the number of cases in Madrid was growing. Altogether, this example highlights the importance of combining mobility and cases to have more broad pictures of the epidemiological process. In the presented example, MAR was calculate by aggregating mobility and cases to the level of provinces. Nevertheless, the MAR score can be calculated at different levels of aggregation. Calculated MAR at different levels of spatial resolution can be download from our systems and it is also included in daily reported individual file in the data repository. The COVID-19 Flow-Board also provides an integrated dashboard for visualising MAR scores between regions at different scales of spatial resolution. The MAR data dashboard can be acceded through the following link: https://flowmaps.life.bsc.es/flowboard-test/board_risk/

**Figure 9.**
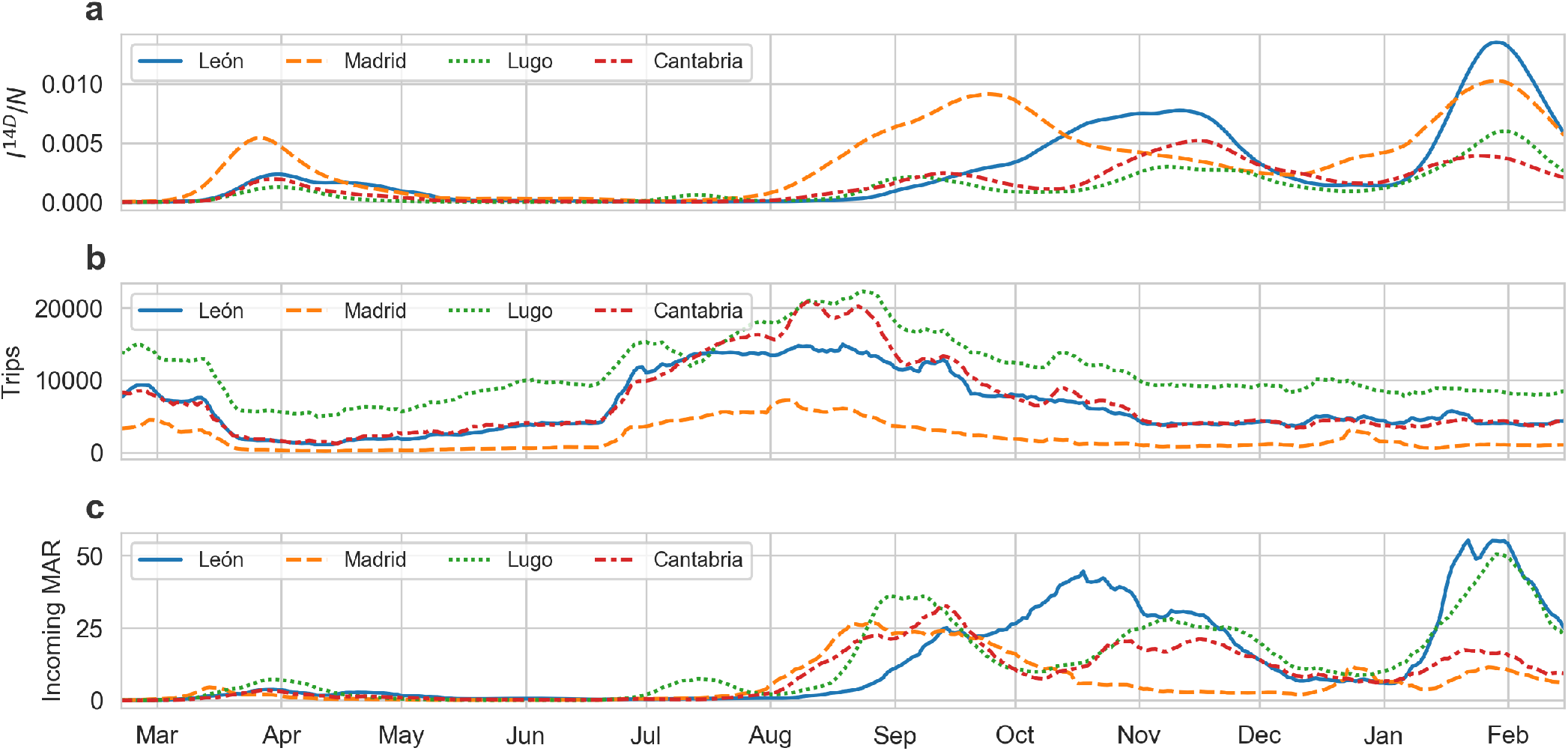
Mobility Associated Risk incoming to Asturias during 2020. The figure shows the time evolution of the COVID-19 cases, the trips and the MAR score for the top four main sources of incoming MAR related to Asturias. Panel (a) shows the evolution of the number of COVID-19 cases per total number of inhabitants. Panel (b) shows the evolution of the trips from the top four sources to Asturias. Panel (c) shows the evolution of MAR incoming to Asturias from the top four main sources of risk.

## Data Availability

All data-sets are available at Zenodo and github. Links and DOIs are included in the main text

https://github.com/bsc-flowmaps

https://zenodo.org/record/4634662

https://zenodo.org/record/4634868

https://zenodo.org/record/4634895

https://zenodo.org/record/4635258

## Code availability

All the code used in the process of data acquisition, processing and analysis have been written in Python Language (version 3.7) and executed in a Linux environment. For data handling, analysis and visualisation we have used the following python libraries: Pandas (0.11.0) SciPy (1.5.2) and Seaborn (0.11.0). All spatial operations have been conducted using GeoPandas (version 0.8.1). The database used to host COVID-19 Flow-Maps is MongoDB (version 4.2.8). The REST-API used to expose the data is implemented using python eve (version 1.1.2). The source code used in this work is available in the public GitHub repository that also hosts the data-set. COVID-19 Flow-Board interactive dashboards were implemented in plotly (version 4.11.0)

### REST-API

We provide a REST API that gives access to the data stored in the MongoDB database. It allows consumers to retrieve multiple documents using query strings, allowing for filtering and sorting. Filters can be provided using MongoDB syntax or Python conditional expressions. It has been created using the open-source python-eve framework (version 1.1.2). Among other fetch-able collections, the API allows downloading the time-dependent population mobility networks across Spain, and daily reports of COVID-19 cases in Spain, at different levels of spatial resolution. The API can be accessed using the endpoint: https://flowmaps.life.bsc.es/api/. Complete documentation with examples can be found at: https://flowmaps.life.bsc.es/api/docs

### Command-line utility and python library for downloading the data

Although all the data records presented in this work is accessible through a REST API, to enable easy access to most relevant collections, we have implemented a lightweight library and a series of scripts in python language to ease the task of downloading the data-sets. The package allows fetching and ingesting the time-dependent population mobility networks across Spain (provided by MITMA), and daily reports of COVID-19 cases in Spain, at different levels of spatial resolution, provided by the National Centre of Epidemiology (CNE) and the different Autonomous Communities. In addition, the package allows downloading the multiple geographical layers for Spain, in GeoJSON format, at the same levels of spatial resolution as the mobility and COVID-19 data. All this code has been packed and published as the standalone python package flowmaps-data. The flowmaps-data toolkit can be installed from pypi repositories with the command pip install flowmaps-data. The package comes with a command line utility that can be used to list, describe and download the data-sets in several formats (CSV, JSON). A complete list of examples and installation instructions can be found here: https://pypi.org/project/flowmaps-data/ Altogether, this toolkit allows the user to query and retrieve different data records including, geographical layer, COVID-19 cases and population mobility, updated on daily basis for Spain.

## Acknowledgements

This work was supported by the Generalitat de Catalunya through the project PDAD14/20/00001, and by the H2020 programme under Grant Agreement 825070 (INFORE) and the INB Grant (PT17/0009/0001 - ISCIII-SGEFI / ERDF).

## Author contributions statement

MPL and AV conceived the study and supervised the project. All authors contributed in finding, curating and annotating data sources. JMF design and implement the data processing workflow. DC, JS, JMF, AV, SCG, JV, MB and MPL participated in design the web-based data dashboards MB and JV developed the web application and code for data accessing. JV implemented the data projecting approaches. TG worked in the processing mobility data. MPL designed and implemented the validation analysis and wrote the first version of draft. All the authors contributed writing and editing the manuscript.

## References

1. Haug, N. et al. Ranking the effectiveness of worldwide COVID-19 government interventions. Nat. Hum. Behav. 4, 1303–1312, 10.1038/s41562-020-01009-0 (2020). Number: 12 Publisher: Nature Publishing Group.

2. Desvars-Larrive, A. et al. A structured open dataset of government interventions in response to COVID-19. Sci. Data 7, 285, 10.1038/s41597-020-00609-9 (2020). Number: 1 Publisher: Nature Publishing Group.

3. Xiong, C., Hu, S., Yang, M., Luo, W. & Zhang, L. Mobile device data reveal the dynamics in a positive relationship between human mobility and covid-19 infections. Proc. Natl. Acad. Sci. 117, 27087–27089 (2020).

4. Badr, H. S. et al. Association between mobility patterns and COVID-19 transmission in the USA: a mathematical modelling study. The Lancet Infect. Dis. 20, 1247–1254, 10.1016/S1473-3099(20)30553-3 (2020). Publisher: Elsevier.

5. Gatto, M. et al. Spread and dynamics of the COVID-19 epidemic in Italy: Effects of emergency containment measures. Proc. Natl. Acad. Sci. 117, 10484–10491, 10.1073/pnas.2004978117 (2020). Publisher: National Academy of Sciences Section: Biological Sciences.

6. Sermi, F, Iacus, S, Vespe, M, Santamaria, C & Spyratos STarchi, D. Mapping Mobility Functional Areas (MFA) using mobile positioning data to inform COVID-19 policies : a European regional analysis. Publ. Off. Eur. Union 10.2760/076318 (2020). ISBN: 9789276204299 Publisher: Publications Office of the European Union.

7. Schlosser, F. et al. COVID-19 lockdown induces disease-mitigating structural changes in mobility networks. Proc. Natl. Acad. Sci. 117, 32883–32890, 10.1073/pnas.2012326117 (2020). Publisher: National Academy of Sciences Section: Physical Sciences.

8. Mitjà, O. et al. Experts request to the Spanish Government: move Spain towards complete lockdown. The Lancet 395, 1193–1194, 10.1016/S0140-6736(20)30753-4 (2020). Publisher: Elsevier.

9. Arenas, A. et al. Modeling the Spatiotemporal Epidemic Spreading of COVID-19 and the Impact of Mobility and Social Distancing Interventions. Phys. Rev. X 10, 041055, 10.1103/PhysRevX.10.041055 (2020). Publisher: American Physical Society.

10. Ferrari, A. et al. Simulating SARS-CoV-2 epidemics by region-specific variables and modeling contact tracing app containment. npj Digit. Medicine 4, 1–8, 10.1038/s41746-020-00374-4 (2021). Number: 1 Publisher: Nature Publishing Group.

11. Kucharski, A. J. et al. Effectiveness of isolation, testing, contact tracing, and physical distancing on reducing transmission of SARS-CoV-2 in different settings: a mathematical modelling study. The Lancet Infect. Dis. 20, 1151–1160, 10.1016/S1473-3099(20)30457-6 (2020). Publisher: Elsevier.

12. Chang, S. et al. Mobility network models of COVID-19 explain inequities and inform reopening. Nature 589, 82–87, 10.1038/s41586-020-2923-3 (2021). Number: 7840 Publisher: Nature Publishing Group.

13. Thompson, R.N. Epidemiological models are important tools for guiding COVID-19 interventions. BMC Medicine 18, 152, 10.1186/s12916-020-01628-4 (2020).

14. Karin, O. et al. Cyclic exit strategies to suppress COVID-19 and allow economic activity. medRxiv 2020.04.04.20053579, 10.1101/2020.04.04.20053579 (2020). Publisher: Cold Spring Harbor Laboratory Press.

15. Vespignani, A. et al. Modelling COVID-19. Nat. Rev. Phys. 2, 279–281, 10.1038/s42254-020-0178-4 (2020). Number: 6 Publisher: Nature Publishing Group.

16. Grantz, K. H. et al. The use of mobile phone data to inform analysis of covid-19 pandemic epidemiology. Nat. communications 11, 1–8 (2020).

17. Pepe, E. et al. COVID-19 outbreak response, a dataset to assess mobility changes in Italy following national lockdown. Sci. Data 7, 230, 10.1038/s41597-020-00575-2 (2020). Number: 1 Publisher: Nature Publishing Group.

18. Kang, Y. et al. Multiscale dynamic human mobility flow dataset in the U.S. during the COVID-19 epidemic. Sci. Data 7, 390, 10.1038/s41597-020-00734-5 (2020). Number: 1 Publisher: Nature Publishing Group.

19. Dong, E., Du, H. & Gardner, L. An interactive web-based dashboard to track COVID-19 in real time. The Lancet Infect. Dis. 20, 533–534, 10.1016/S1473-3099(20)30120-1 (2020). Publisher: Elsevier.

20. Xu, B. et al. Epidemiological data from the COVID-19 outbreak, real-time case information. Sci. Data 7, 106, 10.1038/s41597-020-0448-0 (2020). Number: 1 Publisher: Nature Publishing Group.

21. Ministerio de Transportes, Movilidad y Agenda Urbana (MITMA). Análisis de la movilidad en España con tecnología Big Data durante el estado de alarma para la gestión de la crisis del COVID-19: Informe metodológico (PDF) (2020).

22. European Forum for GeoStatistics. ESSnet project GEOSTAT 1A-Representing Census data in a European population grid-Final Report. (2012).

23. Ministerio de Política Territorial y Función Pública. Registro de entidades locales (2019).

24. Instituto Nacional de Estadistica. https://www.ine.es/en/covid/covid_movilidad_en.htm. Mobility Reduction.

25. News, B. Storm Filomena: Spain sees ’exceptional’ snowfall. BBC News (2021).

